# Quality of life profile of methadone maintenance treatment patients in Ho Chi Minh City, Vietnam

**DOI:** 10.1101/2021.08.06.21261540

**Authors:** Vu Thu Trang, Le Ngoc Tu, Vu Thi Tuong Vi, Khuong Quynh Long, Le Huynh Thi Cam Hong, Tieu Thi Thu Van, Do Van Dung

**Author notes:** These authors contribute equally to this work and act as co-first authors (Vu Thu Trang, Le Ngoc Tu). **Corresponding author:** Vu Thi Tuong Vi, Vietnam HIV Addiction Technology Transfer Center - University of Medicine and Pharmacy at Ho Chi Minh City, Vietnam, 217 Hong Bang Street, Ward 11, District 5, Ho Chi Minh City, Vietnam. **Declarations**. **Ethics approval and consent to participate** All procedures performed in studies involving human participants were in accordance with the ethical standards of The Institution Review Board of Ho Chi Minh City HIV/AIDS Center (IRB-02-2018, dated 10/08/2018). All participants provided signed informed consent. **Consent for publication** Not applicable. **Availability of data and material** Available upon request to the corresponding author. **Conflict of interest** No potential conflict of interest was reported by the authors. **Authors’ contributions** VTT: Conducted data analysis and result explanation, drafted the manuscript. LNT: Collected the data, conducted data analysis and result explanation, drafted the manuscript. KQL: Designed the study, conducted data analysis and result explanation, provided editorial input. VTTV: Designed the study, provided editorial input. LHTCH: Participated in study design, provided editorial input. TTTV: Participated in study design, provided editorial input. DVD: Participated in study design, provided editorial input. All authors read and approved the final manuscript.

## Abstract

**Aim:** To determine the health-related quality of life (HRQoL) of methadone maintenance treatment patients in Ho Chi Minh City, Vietnam.

**Subject and Methods:** A cross-sectional study was conducted in 967 patients treating at two methadone clinics in Ho Chi Minh City, in 2018. Patient’s health-related quality of life was estimated using the EQ-5D-5L and Visual analogue scale (VAS). Tobit regressions were used to identify factors related to patient’s health-related quality of life.

**Results:** Overall, the mean EQ-5D-5L utility and EQ-VAS indexes were 0.96 (SD = 0.12) and 75.8 (SD=15.5), respectively. Factors related with a higher EQ-5D-5L score included peoples who are single, and have a higher monthly income (more than 4 million VND per month), while patients aged under 30 years old, have full-time employment, and have higher education were associated with a higher EQ-VAS score. HIV was associated with lower scores of both EQ-5D-5L and EQ- VAS (β = -0.07 (95%CI: -0.13; -0.01), and β = -7.10 (95%CI: -9.23; -4.98), respectively).

**Conclusion:** HRQoL measurement provides valuable information for the policymaker to adopt suitable decisions on opioid dependence treatment. The finding shows that patients with education, job situation, and socioeconomic status are the related elements with higher HRQoL, which suggested that the policymakers and physicians should pay more attention to these aspects while working on treatment plan for drug users.

## Introduction

Since the first introduction in 1947, Methadone maintenance treatment (MMT) has been proved as an efficacious drug treatment modality for heroin addiction (Herget 2005; O’Donnell and Vogenberg 2011; World Health 2004). In Vietnam, the MMT program was originated in 2008, with approximately 53,000 drug users have received treatment in 336 nationwide MMT clinics up to 31th December 2019 (Nguyen et al. 2017c). Coverage of the MMT program in Vietnam had reached 28% of the total number of people addicted to opiates, treatment adherence rate after six months was 83%, corresponding to the graded good (by World Health Organization standard is 80%). Methadone treatment in Vietnam has been shown to be effective in helping patients reduce and eventually stop using illegal drugs, improve health (reduce HIV infection and diseases transmitted through blood, physical enhancement, physical and mental rehabilitation) (MOH 2020).

Health-related quality of life (HRQoL) is a multidimensional construct related to the physical, mental, emotional, and social functioning of an individual, based on their perceptions. HRQoL is becoming increasingly important in assessing treatment outcomes since most opioid-dependent people often experience negative socioeconomic consequences and social exclusion, let alone suffer from adverse health outcomes and high rates of overdose deaths (Strada et al. 2017). Several studies have found the opioid users or those receiving MMT tend to have impaired HRQoL caused by their comorbid infectious diseases, such as hepatitis B virus (HBV), hepatitis C virus (HCV), and HIV (Astals et al. 2008; Korthuis et al. 2008). For instance, Astals et al. conducted a study to assess the quality of life between the general European population and opioid users, the result found that the former group had significantly lower scores compared to the general population (Astals et al. 2008). Other studies used the patient’s HRQoL as outcome indicator to evaluate the effectiveness of MMT program (Karow et al. 2011; Lashkaripour et al. 2012; Strada et al. 2017). For example, research conducted in HCMC and Hai Phong showed the number of patients with good HRQoL increased from 16% to 55% after 3 months of treatment (Vietnamese Ministry of Health 2014).

Although many studies have explored the HRQoL of opioid users, only a few have examined variations in their scores and determinants factors (Korthuis et al. 2008; Nguyen et al. 2017b; Quyen et al. 2020). A comprehensive understanding of the relationships between HRQoL and MMT is important for developing response strategies to the opioid. This is especially of importance in the context of Vietnam, there are little shreds of evidence that summarize specific linking associated factors to the quality of life for people with opioid dependence. This study was conducted to examine the HRQoL and its determinants among MMT patients in Ho Chi Minh City, Vietnam.

## Method

### Setting and participants

This cross-sectional study was conducted at two MMT clinics in District 8 and District 4, Ho Chi Minh City (HCMC). The inclusion criteria for the survey included: (1) ≥ 18 year olds, (2) currently visiting the clinic during the study period and (3) having the ability to answer the questionnaire. There were 1,039 eligible patients in two clinics, among them, 967 people (93.1%) agreed and participated in the study.

### Procedure and measurement

Patients who met the inclusion criteria were invited to private rooms. Subsequently, they were given the verbal introduction of the study and confirmed their enrollment by signing the consent form. Those who refused to participate continued their usual treatment at the clinic. The questionnaire was conducted by face-to-face interviews in around 25-30 minutes. Data collectors were well-trained preventive medicine doctors from Addiction Technology Transfer Center, University of Medicine and Pharmacy at Ho Chi Minh City. Clinic staffs were not involved in this process to avoid potential bias.

### Outcome variables

The primary outcome of this study was quality of life measured by The EuroQol-five (EQ-5D-5L) and The Visual Analogue Scale (EQ-VAS). The EQ-5D-5L includes five dimensions which describe different aspects of life: mobility, self-care, usual activities, pain/discomfort, and anxiety/depression. In each dimension, a Likert rating scale of five response levels (no problems- code 1, slight problems-code 2, moderate problems-code 3, severe problems- code 4, unable to/extreme problems-code 5) is used. The numbers for each dimension can be combined to a 5- digit code ranging from 11111 (no problems) to 55555 (worst health). The overall EQ-5D-5L score is calculated by using the crosswalk value set in the data of EuroQuoL (EuroQol Research Foundation 2019).

The Visual Analogue Scale (EQ-VAS) is developed to assess the self-rated health of patients. The scale ranges from 0 (*the worst health you can imagine)* to 100 points (*the best health you can imagine*) (EuroQol Research Foundation 2019). The Vietnamese version of EQ-5D-5L and EQ-VAS have been translated, validated and used in many previous studies in Vietnam (Nguyen et al. 2017a; Tran et al. 2018; Tran et al. 2012).

### The Alcohol, Smoking and Substance Involvement Screening Test (ASSIST)

The Alcohol, Smoking and Substance Involvement Screening Test (ASSIST) was included to investigate the risk of concurrent drug use among respondents. The ASSIST was developed by the Word Health Organization (WHO) and designed to be culturally neutral and useable in primary care setting across a variety of cultures. The tool contains a total of 8 questions collecting information about lifetime use of 9 common substances, use of these substances and associated problems over the last 3 months. Patients being assessed as *“medium risk”* or *“high risk”* were defined as hazardous substance users (World Health Organization (WHO) 2010).

### Clinical data extraction

MMT treatment information of participants including duration on MMT, daily methadone dosage, number of doses missed within last 3 months and comorbidities (HIV, tuberculosis, hepatitis B, and hepatitis C) were extracted from the medical records.

Urine samples of respondents were collected and tested for methamphetamine (a substance of ATS group) using rapid urine test (ABON, Biopham Co. Ltd). This process was implemented by well-trained laboratory staffs.

### Data analysis

Descriptive statistics were used to summarize the data, with frequencies and percentages for categorical variables and means with standard deviations (SD) or median with inter-quartile ranges (IQR) for quantitative variables. To detect the differences between EQ-5D-5L and EQ-VAS scores among respondents, we used t-tests, ANOVA tests or Wilcoxon Rank Sum tests when appropriate. Tobit regressions were conducted to identify factors associated with EQ-5D-5L and EQ-VAS scores. A significant level of p < 0.05 was used for all statistical tests. All analyses were carried out using Stata v16 (Stata Corp, College Station, TX).

### Ethics

All procedures performed in studies involving human participants were in accordance with the ethical standards of the Institution Review Board of Ho Chi Minh City HIV/AIDS Center (IRB- 02-2018, dated 10/08/2018). All participants provided signed informed consent.

## Results

A total of 967 participants enrolled into the study. Patients’ demographic characteristics are shown in Table 1. Most of participants were male (89.9%), aged from 30 to 49 (81.8%), having full-time job (74.3%) and being married or living with partners (46.4%). Nearly a haft of respondents (47.8%) finished secondary school and had a monthly income from 4 to 8 million VND (44.8%). The majority of patients had ever injected drugs (78.0%), while only 12.4% respondents still injected drugs within the past three months. About 60% of them had the duration on MMT between 1 to 5 years, with prevalent dose for methadone was 60-120 mg/day (41.2%). Moreover, 44.8% participants reported missing at least one dose in past 3 months. The proportions of participants with positive HIV, HBV, HCV and tuberculosis were 33.9%, 9.7%, 41.9% and 1.0%, respectively.

**Table 1:** Demographic characteristics of respondents.

| Characteristics | Total<br>n | % |
| --- | --- | --- |
| Gender | 967 |  |
| Male | 869 | 89.9 |
| Female | 98 | 10.1 |
| Age | 961 |  |
| < 30 | 111 | 11.6 |
| 30 - 49 | 786 | 81.8 |
| ≥ 50 | 64 | 6.6 |
| Education | 966 |  |
| Primary school or less | 210 | 21.7 |
| Secondary school | 462 | 47.8 |
| High school or above | 294 | 30.4 |
| Occupation | 967 |  |
| Full-time job | 718 | 74.3 |
| Part-time job | 108 | 11.2 |
| Unemployed | 141 | 14.5 |
| Marital status | 966 |  |
| Single | 363 | 37.6 |
| Married/live with partners | 448 | 46.4 |
| Divorced/Separated/ Widowed | 155 | 16.0 |
| Monthly income | 965 |  |
| < 4 million VND | 330 | 34.2 |
| 4 - 8 million VND | 432 | 44.8 |
| > 8 million VND | 203 | 21.0 |
| Ever injected drugs | 967 |  |
| Yes | 754 | 78.0 |
| No | 213 | 22.0 |
| Injecting drug use within the past 3 months | 965 |  |
| Yes | 120 | 12.4 |
| No | 845 | 87.6 |
| Duration on MMT | 965 |  |
| < 1 year | 128 | 13.3 |
| 1-5 years | 580 | 60.1 |
| > 5 years | 257 | 26.6 |
| Daily Methadone dose (mg) | 967 |  |
| < 60 mg/day | 245 | 25.3 |
| 60 - 120 mg/day | 398 | 41.2 |
| >120 mg/day | 324 | 33.5 |
| Dose missed in the last 3 months | 967 |  |
| Yes | 433 | 44.8 |
| No | 534 | 55.2 |
| HIV positive | 967 |  |
| Yes | 328 | 33.9 |
| No | 639 | 66.1 |
| ARV treatment | 312 |  |
| Yes | 302 | 96.8 |
| No | 10 | 3.2 |
| Tuberculosis | 967 |  |
| Yes | 10 | 1.0 |
| No | 957 | 99.0 |
| Hepatitis B | 967 |  |
| Yes | 94 | 9.7 |
| No | 873 | 90.3 |
| Hepatitis C | 967 |  |
| Yes | 405 | 41.9 |
| No | 562 | 58.1 |

Notes: n, number of participants; MMT, methadone maintenance treatment; ARV, antiretroviral treatment.

Table 2 shows profiles of EQ-5D-5L domains according to frequencies of each item response. The highest proportion of respondents reporting any problems was in pain/discomfort domain (14.2%), followed by anxiety/ depression (14.1%), usual activities (7.2%) and mobility (6.7%), the lowest percentage was in Self-care (2.8%).

**Table 2:**
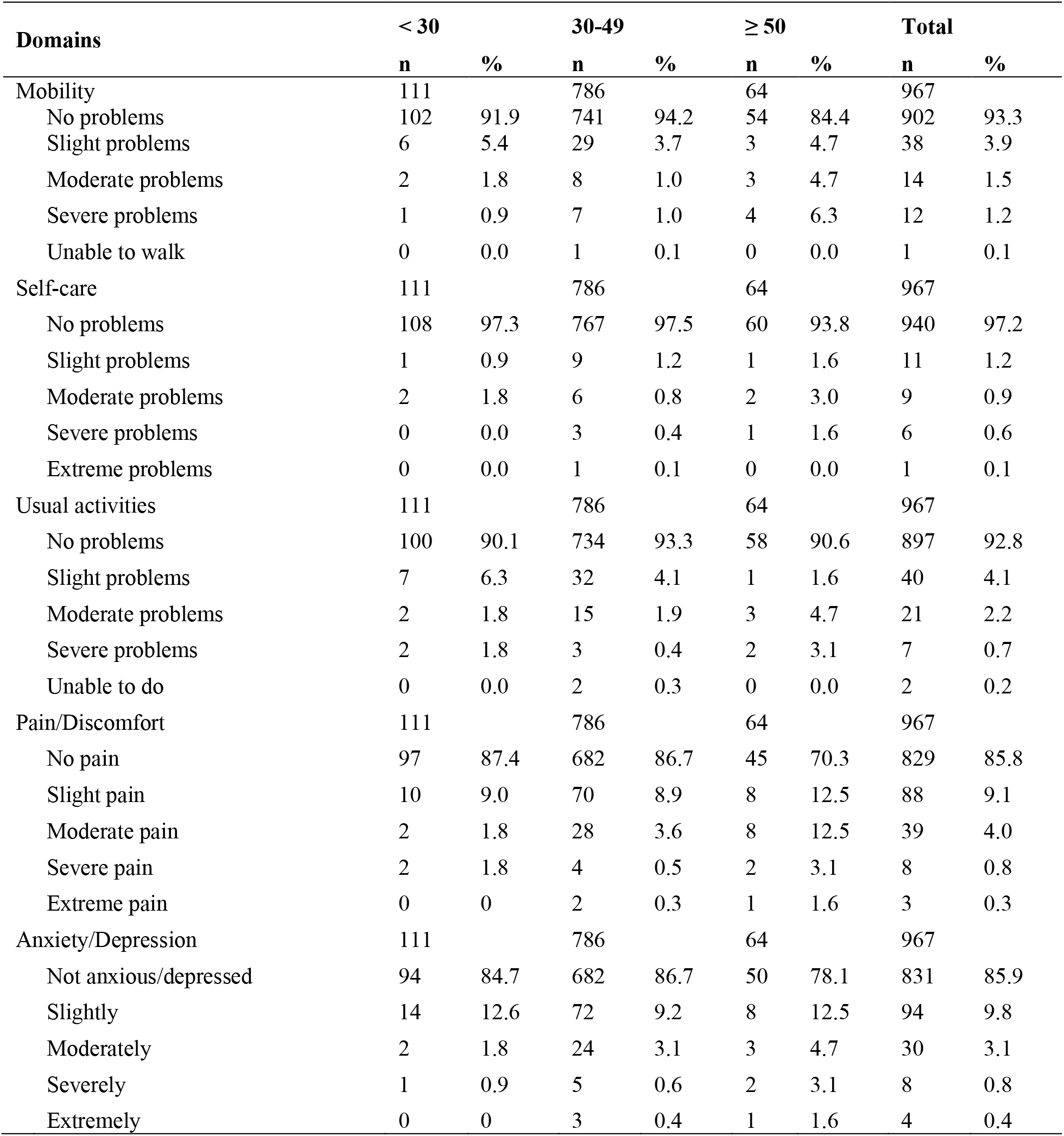
Profiles of EQ-5D-5L by age group.

| Domains | < 30 |  | 30-49 |  | ≥ 50 |  | Total |  |
| --- | --- | --- | --- | --- | --- | --- | --- | --- |
|  | n | % | n | % | n | % | n | % |
| Mobility | 111 |  | 786 |  | 64 |  | 967 |  |
| No problems | 102 | 91.9 | 741 | 94.2 | 54 | 84.4 | 902 | 93.3 |
| Slight problems | 6 | 5.4 | 29 | 3.7 | 3 | 4.7 | 38 | 3.9 |
| Moderate problems | 2 | 1.8 | 8 | 1.0 | 3 | 4.7 | 14 | 1.5 |
| Severe problems | 1 | 0.9 | 7 | 1.0 | 4 | 6.3 | 12 | 1.2 |
| Unable to walk | 0 | 0.0 | 1 | 0.1 | 0 | 0.0 | 1 | 0.1 |
| Self-care | 111 |  | 786 |  | 64 |  | 967 |  |
| No problems | 108 | 97.3 | 767 | 97.5 | 60 | 93.8 | 940 | 97.2 |
| Slight problems | 1 | 0.9 | 9 | 1.2 | 1 | 1.6 | 11 | 1.2 |
| Moderate problems | 2 | 1.8 | 6 | 0.8 | 2 | 3.0 | 9 | 0.9 |
| Severe problems | 0 | 0.0 | 3 | 0.4 | 1 | 1.6 | 6 | 0.6 |
| Extreme problems | 0 | 0.0 | 1 | 0.1 | 0 | 0.0 | 1 | 0.1 |
| Usual activities | 111 |  | 786 |  | 64 |  | 967 |  |
| No problems | 100 | 90.1 | 734 | 93.3 | 58 | 90.6 | 897 | 92.8 |
| Slight problems | 7 | 6.3 | 32 | 4.1 | 1 | 1.6 | 40 | 4.1 |
| Moderate problems | 2 | 1.8 | 15 | 1.9 | 3 | 4.7 | 21 | 2.2 |
| Severe problems | 2 | 1.8 | 3 | 0.4 | 2 | 3.1 | 7 | 0.7 |
| Unable to do | 0 | 0.0 | 2 | 0.3 | 0 | 0.0 | 2 | 0.2 |
| Pain/Discomfort | 111 |  | 786 |  | 64 |  | 967 |  |
| No pain | 97 | 87.4 | 682 | 86.7 | 45 | 70.3 | 829 | 85.8 |
| Slight pain | 10 | 9.0 | 70 | 8.9 | 8 | 12.5 | 88 | 9.1 |
| Moderate pain | 2 | 1.8 | 28 | 3.6 | 8 | 12.5 | 39 | 4.0 |
| Severe pain | 2 | 1.8 | 4 | 0.5 | 2 | 3.1 | 8 | 0.8 |
| Extreme pain | 0 | 0 | 2 | 0.3 | 1 | 1.6 | 3 | 0.3 |
| Anxiety/Depression | 111 |  | 786 |  | 64 |  | 967 |  |
| Not anxious/depressed | 94 | 84.7 | 682 | 86.7 | 50 | 78.1 | 831 | 85.9 |
| Slightly | 14 | 12.6 | 72 | 9.2 | 8 | 12.5 | 94 | 9.8 |
| Moderately | 2 | 1.8 | 24 | 3.1 | 3 | 4.7 | 30 | 3.1 |
| Severely | 1 | 0.9 | 5 | 0.6 | 2 | 3.1 | 8 | 0.8 |
| Extremely | 0 | 0 | 3 | 0.4 | 1 | 1.6 | 4 | 0.4 |

The mean EQ-5D-5L utility scores and EQ-VAS scores by different characteristics are summerized in table 3. Overall, the mean EQ-5D-5L and EQ-VAS indexes were 0.96 (SD=0.12) and 75.8 (SD=15.5), respectively. Lower EQ-5D-5L and EQ-VAS scores were found older patients and those having lower education, and having unstable job. Patients who had monthly income under 4 million VND had lower EQ-5D-5L and EQ-VAS indexs compared to other groups (p<0.001). We also found significant differences on EQ-5D-5L and EQ-VAS scores in term of duration on MMT, daily methadone dose, dose missed in the last 3 months. Notably, people being positive with HIV, hepatitis B, C or having ARV treatment also had lower EQ-5D-5L score or EQ-VAS index than their counterparts.

**Table 3:** EQ-5D-5L utility scores and EQ-VAS scores by different characteristics.

|  | EQ-5D-5L scores |  |  | EQ-VAS scores |  |  |
| --- | --- | --- | --- | --- | --- | --- |
|  | Mean | SD | p-value | Mean | SD | p-value |
| Total | 0.96 | 0.12 |  | 75.8 | 15.5 |  |
| Gender |  |  |  |  |  |  |
| Male | 0.96 | 0.11 | 0.249 | 75.8 | 15.2 | 0.949 |
| Female | 0.94 | 0.19 |  | 75.7 | 17.7 |  |
| Age |  |  |  |  |  |  |
| < 30 | 0.96 | 0.11 | <0.001 | 79.9 | 15.5 | <0.001 |
| 30 - 49 | 0.96 | 0.11 |  | 75.9 | 15.1 |  |
| ≥ 50 | 0.9 | 0.2 |  | 67.3 | 16.6 |  |
| Education |  |  |  |  |  |  |
| Primary school or less | 0.93 | 0.18 | <0.001 | 72.2 | 17.1 | <0.001 |
| Secondary school | 0.96 | 0.11 |  | 76.2 | 15.7 |  |
| High school or above | 0.97 | 0.07 |  | 77.6 | 13.4 |  |
| Occupation |  |  |  |  |  |  |
| Full-time job | 0.97 | 0.1 | <0.001 | 77.5 | 14.4 | <0.001 |
| Part-time job | 0.94 | 0.12 |  | 71.4 | 15.5 |  |
| Unemployed | 0.91 | 0.19 |  | 70.5 | 18.6 |  |
| Marital status |  |  |  |  |  |  |
| Single | 0.97 | 0.09 | 0.058 | 76.2 | 15.1 | 0.791 |
| Married/live with partners | 0.95 | 0.13 |  | 75.6 | 15.3 |  |
| Divorced/Separated/<br>Widowed | 0.94 | 0.15 |  | 75.3 | 16.9 |  |
| Monthly income |  |  |  |  |  |  |
| < 4 million VND | 0.93 | 0.17 | <0.001 | 72.2 | 17.6 | <0.001 |
| 4 - 8 million VND | 0.97 | 0.08 |  | 77.1 | 14.1 |  |
| > 8 million VND | 0.97 | 0.08 |  | 79 | 13.5 |  |
| Ever injected drugs |  |  |  |  |  |  |
| Yes | 0.96 | 0.12 | 0.62 | 75 | 15.6 | 0.005 |
| No | 0.96 | 0.12 |  | 78.4 | 14.9 |  |
| Injecting drug use within the past<br>3 months |  |  |  |  |  |  |
| Yes | 0.95 | 0.09 | 0.818 | 73.3 | 16.7 | 0.059 |
| No | 0.96 | 0.12 |  | 76.1 | 15.3 |  |
| Duration on MMT |  |  |  |  |  |  |
| < 1 year | 0.97 | 0.09 | 0.645 | 77.6 | 14.7 | 0.01 |
| 1-5 years | 0.95 | 0.13 |  | 76.5 | 15.5 |  |
| > 5 years | 0.96 | 0.11 |  | 73.4 | 15.7 |  |
| Daily Methadone dose (mg) |  |  |  |  |  |  |
|  | Mean | SD | p-value | Mean | SD | p-value |
| < 60 mg/day | 0.96 | 0.10 | 0.138 | 77.8 | 14.8 | <0.001 |
| 60 - 120 mg/day | 0.96 | 0.10 |  | 76.7 | 15.5 |  |
| >120 mg/day | 0.95 | 0.15 |  | 73.2 | 15.6 |  |
| Dose missed in the last 3 months |  |  |  |  |  |  |
| Yes | 0.97 | 0.1 | 0.047 | 76.4 | 14.5 | 0.271 |
| No | 0.95 | 0.13 |  | 75.3 | 16.2 |  |
| HIV positive |  |  |  |  |  |  |
| Yes | 0.94 | 0.15 | 0.017 | 71.1 | 15.8 | <0.001 |
| No | 0.96 | 0.1 |  | 78.2 | 14.8 |  |
| ARV treatment (n=312) | 0.94 | 0.15 |  |  |  |  |
| Yes | 0.94 | 0.15 | <0.001 | 70.3 | 15.7 | 0.012 |
| No | 0.99 | 0.03 |  | 83 | 10.3 |  |
| Tuberculosis |  |  |  |  |  |  |
| Yes | 0.9 | 0.31 | 0.589 | 74.5 | 13 | 0.791 |
| No | 0.95 | 0.12 |  | 75.8 | 15.5 |  |
| Hepatitis B |  |  |  |  |  |  |
| Yes | 0.94 | 0.16 | 0.315 | 72 | 15.2 | 0.013 |
| No | 0.96 | 0.11 |  | 76.2 | 15.5 |  |
| Hepatitis C |  |  |  |  |  |  |
| Yes | 0.96 | 0.12 | 0.529 | 74.3 | 15.6 | 0.013 |
| No | 0.95 | 0.12 |  | 76.8 | 15.3 |  |

Table 4 shows the association between EQ-5D-5L and EQ-VAS scores and different kinds of substance use. Patients being hazardous ATS and cannabis users had 5 points and 7 points lower than the others in EQ-VAS index, respectively (p<0.001 for ATS use and p=0.043 for cannabis use).

**Table 4:** EQ-5D-5L utility scores and EQ-VAS scores by different substance uses.

|  | EQ-5D-5L scores |  |  | EQ-VAS scores |  |  |
| --- | --- | --- | --- | --- | --- | --- |
|  | Mean | SD | p-value | Mean | SD | p-value |
| Hazardous tobacco use (ASSIST $\geq 4$) | | | | | | |
| Yes (n=941) | 0.96 | 0.11 | 0.418 | 75.8 | 15.5 | 0.954 |
| No (n=26) | 0.91 | 0.31 |  | 76.0 | 14.3 |  |
| Hazardous alcohol use (ASSIST $\geq 11$) | | | | | | |
| Yes (n=81) | 0.95 | 0.09 | 0.707 | 76.0 | 15.2 | 0.888 |
| No (n=886) | 0.96 | 0.12 |  | 75.8 | 15.5 |  |
| Hazardous ATS use (ASSIST $\geq 4$) | | | | | | |
| Yes (n=161) | 0.95 | 0.13 | 0.251 | 71.7 | 17.0 | **<0.001** |
| No (n=806) | 0.96 | 0.12 |  | 76.6 | 15.1 |  |
| Positive urine test with ATS |  |  |  |  |  |  |
| Yes (n=246) | 0.96 | 0.11 | 0.491 | 76.4 | 14.7 | 0.490 |
| No (n=721) | 0.96 | 0.12 |  | 75.6 | 15.8 |  |
| Hazardous opioids use (ASSIST $\geq 4$) | | | | | | |
| Yes (n=825) | 0.96 | 0.12 | 0.171 | 75.7 | 15.5 | 0.564 |
| No (n=142) | 0.94 | 0.14 |  | 76.5 | 15.5 |  |
| Hazardous cannabis use (ASSIST $\geq 4$) | | | | | | |
| Yes (n=25) | 0.91 | 0.16 | 0.112 | 69.6 | 13.7 | **0.043** |
| No (n=942) | 0.96 | 0.12 |  | 76.0 | 15.5 |  |

Notes: ASSIST, The Alcohol, Smoking and Substance Involvement Screening Test

Table 5 illustrates ten most common EQ-5D-5L health states, which accounted for 91.3% of participants. Health conditions “11111” (full health), “11112” (slightly problems in anxiety/depression) and “11121” (slightly problems in pain/discomfort) were the most frequent statuses among respondents.

**Table 5:** Most frequent EQ-5D-5 L health states with mean utility scores and EQVAS scores.

| Health states | Number | Percent | Cum% | Mean Utility | Mean VAS |
| --- | --- | --- | --- | --- | --- |
| 11111 | 752 | 77.8 | 77.8 | 1.00 | 78.5 |
| 11112 | 37 | 3.8 | 81.6 | 0.94 | 74.6 |
| 11121 | 28 | 2.9 | 84.5 | 0.92 | 65.5 |
| 11122 | 20 | 2.1 | 86.6 | 0.85 | 72.4 |
| 11131 | 16 | 1.7 | 88.3 | 0.85 | 55.6 |
| 11211 | 7 | 0.7 | 89.0 | 0.95 | 71.4 |
| 11113 | 7 | 0.7 | 89.7 | 0.89 | 76.4 |
| 21111 | 6 | 0.6 | 90.3 | 0.93 | 80.0 |
| 11222 | 5 | 0.5 | 90.8 | 0.81 | 70.0 |
| 11221 | 5 | 0.5 | 91.3 | 0.87 | 67.0 |

The final multivariate model revealed four factors related to EQ-5D-5L scores. Patients who were married/live with partners and those who were divorced/separated/widowed had lower EQ-5D-5L scores, compared with patients who were single, (β=-0.08; 95%CI: -0.14,-0.01 and β=-0.09; 95%CI: -0.17,-0.01, respectively). Similarly, the lower scores were found in patients who had unstable job compared to those having full-time job (β=-0.10; 95%CI: -0.19,-0.01 in part-time job and β=-0.09; 95%CI:-0.18,-0.01 in unemployed) or being positive with HIV (β=-0.07; 95%CI: - 0.13,-0.01). By contrast, patients had higher EQ-5D-5L score when they got higher monthly income (β=0.12; 95%CI: 0.04,0.20 in 4-8 million VND and β=0.12; 95%CI: 0.02,0.21 in >8 million VND).

Similar results were found in the final model for EQ-VAS scores. The lower EQ-VAS scores were found in patients having unstable job as compared to those having full-time job (β=-4.24; 95%CI: -7.38,-1.08 for part-time job and β=-5.17; 95%CI: -7.98,-2.36 for unemployed), having HIV positive serostatus (β=-7.10; 95%CI: -9.23,-4.98), aged ≥ 50 years (β=-12.78; 95%CI: -17.56,- 8.01) and having hazardous ATS use (β=-5.97; 95%CI: -8.60,-3.35). Whereas, patients who finished secondary school or above had higher EQ-VAS scores than those who did not. (Table 6)

**Table 6:** Multivariate linear regression model of factors related to EQ-5D-5L and EQ-VAS scores.

|  | EQ-5D-5L scores |  |  | EQ-VAS scores |  |  |
| --- | --- | --- | --- | --- | --- | --- |
| | $\beta$ | 95%CI | p-value | $\beta$ | 95%CI | p-value |
| **Age** | - | - | - |  |  |  |
| < 30 |  |  |  | 1 |  |  |
| 30 - 49 |  |  |  | -2.45 | [-5.59; 0.69] | 0.126 |
| $\geq 50$ | | | | -12.78 | [-17.56; -8.01] | <0.001 |
| **Education** | - | - | - |  |  |  |
| Primary school or less |  |  |  | 1 |  |  |
| Secondary school |  |  |  | 2.92 | [0.41; 5.43] | 0.023 |
| High school or above |  |  |  | 3.71 | [0.98; 6.45] | 0.008 |
| **Marital status** |  |  |  |  |  |  |
| Single | 1 |  |  |  |  |  |
| Married/live with partners | -0.08 | [-0.14; -0.01] | 0.021 |  |  |  |
| Divorced/Separated/ Widowed | -0.09 | [-0.17; -0.01] | 0.045 |  |  |  |
| **Occupation** |  |  |  |  |  |  |
| Full-time job | 1 |  |  | 1 |  |  |
| Part-time job | -0.10 | [-0.19; -0.01] | 0.031 | -4.24 | [-7.38; -1.08] | 0.008 |
| Unemployed | -0.09 | [-0.18; -0.01] | 0.066 | -5.17 | [-7.98; -2.36] | <0.001 |
| **Monthly income** |  |  |  | - | - | - |
| < 4 million VND | 1 |  |  |  |  |  |
| 4 - 8 million VND | 0.12 | [0.04; 0.20] | 0.003 |  |  |  |
| > 8 million VND | 0.12 | [0.02; 0.21] | 0.013 |  |  |  |
| **HIV positive** |  |  |  |  |  |  |
| No | 1 |  |  | 1 |  |  |
| Yes | -0.07 | [-0.13; -0.01] | 0.036 | -7.10 | [-9.23; -4.98] | <0.001 |
| **Hazardous ATS use** | - | - | - |  |  |  |
| No |  |  |  | 1 |  |  |
| Yes |  |  |  | -5.97 | [-8.60; -3.35] | <0.001 |

Notes: CI, confidence interval

## Discussion

The mean score of EQ-5D-5L and EQ-VAS in our study were 0.96 (SD = 0.12) and 75.8 (SD=15.5), respectively, which were higher than that of the methadone patients in the North mountain area in Vietnam 0.88 (SD = 0.20) and long-term treatment with methadone patients in Norway 0.699 (0.25) (Nguyen et al. 2017b). This difference could happen due to the differences in geographical location, social distinctions, economic status, and culture.

Our results indicated that MMT patients aged from 30 to 50 years old usually did not experience significant pain/discomfort and anxiety/depression symptoms in their daily life. These symptoms were more frequently reported among patients who were lower than 30 or older than 50 years old. This finding was consistent with the previous study in Vietnam, in which, the prevalence of anxiety/depression was the most common symptom among study participants (Tran et al. 2011). The two problems (pain/discomfort and anxiety/depression) were also the most frequently reported by MMT patients in Poland (Golicki and Niewada 2017). Notably, 77.8% of participants in our study reported with the health states as 11111 (perfect health state) indicated that the quality of life among MMT patients in Vietnam was relatively high.

We found that the patient’s age, education, marital status, employment status, and socioeconomic status were significantly associated with their HRQoL. Specifically, the HRQoL scores were lower among older patients, which is in line with previous research in Vietnam, Uruguay, England and Germany (Augustovski et al. 2016; Feng et al. 2015; Hinz et al. 2014). Patients with higher education, being employed, and having high socioeconomic had higher HRQoL than the other groups. It could be explained that people with stable income and jobs are easier to access MMT services, and people with higher education are more likely to adopt/adherence the MMT treatment and have the self-awareness to break the addiction (Quyen et al. 2020; Sadeghi et al. 2017).

Consistent with previous studies, our study revealed patients involving in the MMT program more than five years had lower score of EQ-VAS as compared to those being treated in less than 5 years. (Babaie and Razeghi 2013; Quyen et al. 2020). The results also showed patients who missed dose had a higher score than adherence patients. Since the causal relationship has not been confirmed yet, this finding raises the question that MMT treatment effective may lead to miss dose decision in MMT patient.

In the present study, 33.9% of participants had HIV, and 96.8% of them were on ARV treatment, among these, we found patients living with HIV had lower HRQoL. According to previous studies, comorbidities contribute to lower HRQoL because they lead to poorer health status (Tran 2012; Tran et al. 2011; Wang et al. 2014). ARVs have the effect of inhibiting the reproduction of HIV but can bring side effects for the users. Therefore, it is as expected that the lower HRQoL scores were found among these patients. However, the most difficult thing that these patients had to face was experiencing social stigma and discrimination (Quyen et al. 2020).

Besides, HIV patients also bear the burden of treatment-related costs and are limited in daily activities (Wang et al. 2014). In Vietnam, health insurance does not cover the costs of daily consumables that methadone used but pays for routine laboratory tests and costs for treatment of adverse effects and comorbidities in compliance with regulations by Vietnam’s Ministry of Health. Accordingly, drug users with insurance when participating in MMT are less worried about paying additional treatment and laboratory test costs. The previous study has shown this policy helps insurance patients sought and received treatment more promptly, sufficiently, and properly than those who had to pay for treatment by themselves (Quyen et al. 2020).

In our study, patients being hazardous ATS users had lower EQ-VAS scores than their counterparts. A previous study demonstrated that drug use among MMT patients was still a rising problem that could decrease their health and quality of life (Le et al. 2021). Long-term ATS use negatively leads to intoxication, emotional disorder, anxiety and increases high-risk sexual behaviors such as unprotected sex and sex with multiple partners (Radfar and Rawson 2014; Volkow et al. 2007). Our finding indicates the need for programs on controlling ATS use among MMT patients, including screening, provide urine testing for the patient as a regular test. Besides, other interventions to help patients withdraw from ATS are needed to maintain better outcomes for MMT patients (Shariatirad et al. 2013).

Our study has several limitations. First, as a cross-sectional survey, the results could not be interpreted as causal relationship. Second, as all the participants are recruited in Ho Chi Minh City, the results might not be generalized for MMT patients in other areas. Further studies in Vietnam are needed to thoroughly explore the associated factors with HRQoL among MMT patients, including the different elements between male and female patients, social stigma, or psychology events that could affect HRQoL.

## Conclusion

HRQoL measurement provides valuable information for the policymaker to adopt suitable decisions on opioid dependence treatment. The finding shows that patients with education, job situation, and socioeconomic status are the related elements with higher HRQoL, which suggested that the policymakers and physicians should pay more attention to these aspects while working on treatment plan for drug users.

## Data Availability

Due to the nature of this research, participants of this study did not agree for their data to be shared publicly, so supporting data is not available.

## Acknowledgements

The authors would like to thank all MMT patients who participated in this study, colleagues in the South Vietnam HIV Addiction Technology Transfer Center and Ho Chi Minh City HIV/AIDS Center, District 4 and District 8 MMT clinics for supporting this research.

